# Correlation between continuous Positive end-expiratory pressure (PEEP) values and occurrence of Pneumothorax and Pneumomediastinum in SARS-CoV2 patients during non-invasive ventilation with Helmet

**DOI:** 10.1101/2020.08.31.20185348

**Authors:** Antonio Gidaro, Federica Samartin, Anna M. Brambilla, Chiara Cogliati, Stella Ingrassia, Francesco Banfi, Viola Cupiraggi, Cecilia Bonino, Marco Schiuma, Andrea Giacomelli, Stefano Rusconi, Jaqueline Currà, Antonio Luca Brucato, Emanuele Salvi

## Abstract

**Background:** Acute Hypoxemic Respiratory Failure is a common complication of SARS-CoV2 related pneumonia, for which non-invasive ventilation (NIV) with Helmet Continuous Positive Airway Pressure (CPAP) is widely used. The frequency of pneumothorax in SARS-CoV2 was reported in 0.95% of hospitalized patients in 6% of mechanically ventilated patients, and in 1% of a post-mortem case series.

**Objectives:** Aim of our retrospective study was to investigate the incidence of pneumothorax and pneumomediastinum (PNX/PNM) in SARS-CoV2 pneumonia patients treated with Helmet CPAP. Moreover, we examined the correlation between PNX/PNM and Positive end-expiratory pressure (PEEP) values.

**Methods:** We collected data from patients admitted to “Luigi Sacco” University Hospital of Milan from 2 February to 5 May 2020 with SARS-CoV2 pneumonia requiring CPAP. Patients, who need NIV with bi-level pressure or endotracheal intubation (ETI) for any reason except those who needed ETI after PNX/PNM, were excluded. Population was divided in two groups according to PEEP level used (≤10 cmH2O and >10 cmH20).

**Results:** 154 patients were enrolled. In the overall population, 42 patients (27%) were treated with High-PEEP (>10 cmH2O), and 112 with Low-PEEP (≤10 cmH2O). During hospitalization 3 PNX and 2 PNM occurred (3.2%). Out of these five patients, 2 needed invasive ventilation after PNX and died. All the PNX/PNM occurred in the High-PEEP group (5/37 vs 0/112, p<0,001).

**Conclusion:** The incidence of PNX appears to be lower in SARS-CoV2 than SARS and MERS. Considering the association of PNX/PNM with high PEEP we suggest using the lower PEEP as possible to prevent these complications.

## INTRODUCTION

Coronavirus disease 2019 (COVID-19) is an infectious disease caused by severe acute respiratory syndrome coronavirus 2 (SARS-CoV-2) which may cause life-threatening diseases (with dyspnoea, hypoxia, or >50 percent lung involvement on imaging within 24 to 48 hours) in 14% of patients, even leading to critical diseases in 5%^1^: Acute Hypoxemic Respiratory Failure (AHRF), defined as severe arterial hypoxemia (pO2<60 mmHg), refractory to supplemental oxygen, and Acute Respiratory Distress Syndrome (ARDS), defined by a PaO2/FiO2 ratio less than 300 mmHg with PEEP ≥ 5 cm H2O, according to the Berlin Definition.

During this novel coronavirus pandemic, non-invasive ventilation (NIV) and Continuous Positive Airway Pressure (CPAP) were widely used to treat AHRF gaining respiratory support^2^. NIV has already been used during SARS and MERS epidemic with some evidence of efficacy^3-5^ and previous reports support its use in pneumonia^6,7^. Moreover, during SARS-CoV2 outbreak the overcrowding of the ICUs pushed to use a bridge or alternative respiratory supports in medical wards. During SARS epidemic in 2002, NIV was used to treat acute respiratory failure in SARS pneumoniae with different devices: Helmet CPAP, Nasal CPAP, BiPaP^3,4^. Between many interfaces available for CPAP therapy, the helmet has been proposed to reduce droplet dispersion and consequently preventing health care worker’s infection^8^ during SARS-CoV2 pandemic and, for the same reason, the antimicrobial filter was adopted. During the past epidemics, 4 to 15 cmH2O^3,4^ of positive end-expiratory pressure (PEEP) were administered and higher PEEP were contraindicated because of the elevated incidence of pneumomediastinum (PNM) and pneumothorax (PNX) in patients with SARS and MERS pneumonia (1.7-12%^9,10^ and 16.4%^11^ respectively, either spontaneous or associated to ventilation). Conversely, these complications have not been reported when NIV was used for the treatment of common pneumonia patients^6,7^.

For SARS-CoV2 so far, the frequency of pneumothorax has been reported in 0.95% of hospitalized patients^12^, in 6% of mechanically ventilated patients^13^, and in 1% of a post-mortem case series^14^. The first aim of our study was to 1) assess the incidence of PNX and PNM in SARS-CoV2 pneumonia patients treated with Helmet CPAP; 2) investigate the correlation between the incidence of PNX or PNM and the PEEP values used during the treatment.

## MATERIALS AND METHODS

### Study design and setting

We retrospectively analysed data from patients admitted in Hospital Wards of “Luigi Sacco” University Hospital of Milan from the Emergency Department (ER) or transferred from other hospitals of Lombardy region from 2^nd^ February to 5^th^ May 2020.

The inclusion criteria were: 1) Chest X-Ray positive for bilateral pneumonia SARS-CoV-2 related. SARS-CoV-2 infection was defined by the positivity of real-time reverse transcription-polymerase chain reaction test (RT-PCR) at nose-pharyngeal swab with pneumonia 2) acute hypoxemic respiratory failure (PaO2/FIO2 ≤ 150 mm Hg and respiratory rate>25/minute) requiring helmet CPAP treatment.

The exclusion criterion was the need of NIV with bi-level pressure not performable through (Helmet) and endotracheal intubation (ETI) for any reason except for those who needed ETI after PNX/PNM.

PNX and PNM were documented with chest X-ray, usually carried out for worsening of clinical conditions.

The study (“REGISTRO DELLE INFEZIONI SOSPETTE E ACCERTATE COVID-19/Studio Sacco COVID-19)” was approved by the local ethical committee with the registration number 2020/16088.

### Statistical Analysis

We divided the index population in two groups according to PEEP level used under CPAP (≤10 cmH2O and >10 cmH20).

Kolmogorov-Smirnov test was done to evaluate the normality of distribution of data. Qualitative data were expressed as number and percentage. Chi square or Fisher exact tests were used in group’s comparison. Quantitative data were expressed as mean, standard deviation, median and range. Student T-test and Mann-Whitney test (for non-parametric data) were used for comparison between groups. *P*-value less than 0.05 was considered statistically significant.

The statistical analysis of data was done by using ***Excel*** (Office program 2016) and ***SPSS*** (statistical package for social science-SPSS, Inc., Chicago, IL version 20).

## RESULTS

During the observational period, 1016 patients were admitted to our Hospital, 194 (19.1%) met the inclusion criteria. Of them, 40 patients (3,6%) were excluded because they needed ETI, reducing the eligible number to 154 (15.2%).

Table 1 summarizes the clinical characteristics of the examined population; the average age was 68.8 years (± 14.7; range 27-94 years), 107 were men (69.5%) and 47 women (30.5%). CPAP was performed with PEEP set up between 5 to 15 cmH2O (modal value 10 cmH2O) and FiO2 from 30% to 100%. The average duration of CPAP treatment was 174 hours – 6,4 days – with a great heterogeneity (± 141; range 3-754 hour). Mortality rate for the whole population was 25.3%. During the observational period, 3 PNX and 2 PNM events occurred (total 5 events). Between these cases, no one had a prior story of smoking or underlying lung disease and none of them required thoracic invasive procedures (as positioning central venous catheter or thoracentesis). The most relevant features of these five patients are reported in Table 2.

**Table 1.** Characteristic of examined population and the 2 groups according to PEEP value Continuous variables are expressed as mean ± standard deviation

|  | GENERAL POPULATION | LOW PEEP GROUP | HIGH PEEP GROUP | P VALUE |
| --- | --- | --- | --- | --- |
| Number of patients | 154 | 112 | 42 |  |
| Age (years) | 65.8 ± 14.7 | 65.8 ± 14.5 | 66.0 ± 15.4 | 0.9 |
| Male Sex | 107 (69.5%) | 73 (65.2%) | 34 (80.9%) | 0.058 |
| PN<sub>X</sub>/PN<sub>M</sub> | 5 (3.2%) | 0 (0%) | 5 (11.9%) | **<0.001** |
| Deaths | 39 (25.3%) | 24 (21.4%) | 15 (35.7%) | 0.069 |
| Tocilizumab | 50 (32.5%) | 33 (29.5%) | 17 (40.5%) | 0.183 |
| High dose steroids | 16 (10.4%) | 12 (10.7%) | 4 (9.5%) | 0.829 |
| RR at T0 (bpm)* | 27 ± 7 | 26 ± 7 | 27 ± 8 | 0.63 |
| pO<sub>2</sub> at T0 (mmHg)<sup>¥</sup> | 68.3 ± 21.8 | 67.9 ± 22.0 | 69.2 ± 21.6 | 0.74 |
| pH at T0 | 7.47 ± 0.06 | 7.47 ± 0.06 | 7.46 ± 0.07 | 0.32 |
| FiO<sub>2</sub> at T0 (%) | 40.3 ± 25.3 | 37.7 ± 23.9 | 47.1 ± 27.83 | 0.058 |
| pO<sub>2</sub>/FiO<sub>2</sub> at T0 | 223 ± 100 | 232 ± 96 | 202 ± 108 | 0.1 |
| Length of hospitalization (days) | 20.3 ± 14.4 | 20.8 ± 15.0 | 19.1 ± 13.0 | 0.52 |
| Duration of CPAP (hours) | 174.5 ± 140.8 | 177.3 ± 150.0 | 166.8 ± 113.7 | 0.68 |
| Duration of symptoms (days) | 7.3 ± 3.9 | 7.1 ± 3.9 | 7.6 ± 3.9 | 0.49 |
| CRP (mg/L)<sup>⊙</sup> | 159 ± 92 | 158 ± 93 | 161 ± 92 | 0.85 |
| IL-6 | 144 ± 412 | 103 ± 211 | 246 ± 687 | 0.248 |
| GRs/Ly ratio<sup>⊠</sup> | 8.1 ± 7.1 | 7.7 ± 5.7 | 9.1 ± 9.9 | 0.28 |

T0 = before starting CPAP
\* Respiratory rate before starting CPAP
¥ Partial pressure of oxygen in arterial blood
⊠ Partial pressure of carbon dioxide in arterial blood
^ Oxygen saturation in arterial blood
~ Charlson Comorbidity Index
⊙ C-Reactive Protein
⊠ Granulocytes/Lymphocytes ratio

**Table 2.** Clinical and biochemical features about 5 cases of PNX/PNM

|  | CASE 1 | CASE 2 | CASE 3 | CASE 4 | CASE 5 |
| --- | --- | --- | --- | --- | --- |
| AGE (RANGE) | 40's | 70's | 40's | 70's | 70's |
| SEX | Male | Female | Male | Male | Female |
| SMOKING HISTORY | No | No | No | No | No |
| CHARLSON INDEX | 0 | 3 | 0 | 4 | 3 |
| UNDERLYING LUNG DISEASE | No | No | No | No | No |
| CHEST RX FINDINGS ON HOSPITAL ADMISSION | Right upper lobe and bilateral lower | Left upper lobe and right lower lobe | Bilateral all lobe | Bilateral all lobe | Bilateral medium and low lobe |
| PEAK LDH (U/L) | 313 | 662 | 671 | 507 | 634 |

**Table 2.** Clinical and biochemical features about 5 cases of PNx/PMN
|  |  |  |  |  |  |
| --- | --- | --- | --- | --- | --- |
| **PEAK PCR (MG/L)** | 203 | 209 | 142 | 260 | 372 |
| **PEAK IL6 (NG/L)** | 76 | 65 | 60 | 105 | Not Performed |
| **PEAK D-DIMER (MG/L FEU)** | 426 | 3443 | 795 | 5985 | 3213 |
| **LOWEST LYMPHOCYTE COUNT (10<sup>6</sup>/L)** | 860 | 640 | 1200 | 750 | 330 |
| **TIME FROM BEGINNING OF SYMPTOMS AND PNx (DAYS)** | 17 | 23 | 9 | 23 | 10 |
| **TIME BETWEEN STARTING CPAP AND PNx/PMN (HOURS)** | 168 | 170 | 72 | 408 | 72 |
| **PEEP CPAP (CMH2O)** | 12,5 | 15 | 12,5 | 12,5 | 12,5 |
| **HIGHER FIO2 DURING CPAP** | 60% | 60% | 60% | 60% | 50% |
| **HIGHER FIO2 AFTER PNx/PMN** | 100% | 100% | 100% | 100% | 100% |
| **SIDE OF PNx** | Bilateral | No | Left lung | No | Bilateral |
| **PNM** | No | Yes | No | Yes | Yes |
| **ANTIVIRAL THERAPY** | Ritonavir/Lopinavir | Ritonavir/Lopinavir + Remdesivir | Ritonavir/Lopinavir | Ritonavir/Lopinavir + Remdesivir | Ritonavir/Lopinavir + Remdesivir |
| **USE OF STEROID** | No | Yes | No | Yes | No |
| **USE OF TOCILIZUMAB** | Yes | Yes | No | No | No |
| **REQUIRED ETI AFTER PNx** | No | No | Yes | No | Yes |
| **CHEST DRAINAGE AFTER PNx** | No | No | Yes | No | Yes |
| **TIME FROM PNx AND RESOLUTION OF PNx/CHEST TUBE REMOVAL** | 8 | 10 | 5 | 12 | 15 |
| **TIME FROM PNx AND DISCHARGE/DEATH (DAYS)** | 18 | 20 | 5 | 54 | 25 |
| **DEATH** | No | No | Yes | No | Yes |

After PNX or PNM events, every patient needed higher FiO2 (from 50-60% to 100%): 2 underwent chest drainage and subsequently ETI (Figure 1) and then died (mortality rate of 40% of this small subset). The average time elapsed from starting CPAP to the occurrence of PNX/PMN was 180 hours with great heterogeneity (±137; range 72-408 hour). No difference was found in the duration of CPAP treatment between PNX/PMN group and No-PNX/PMN patients (p-value 0.931).

**Figure 1:**
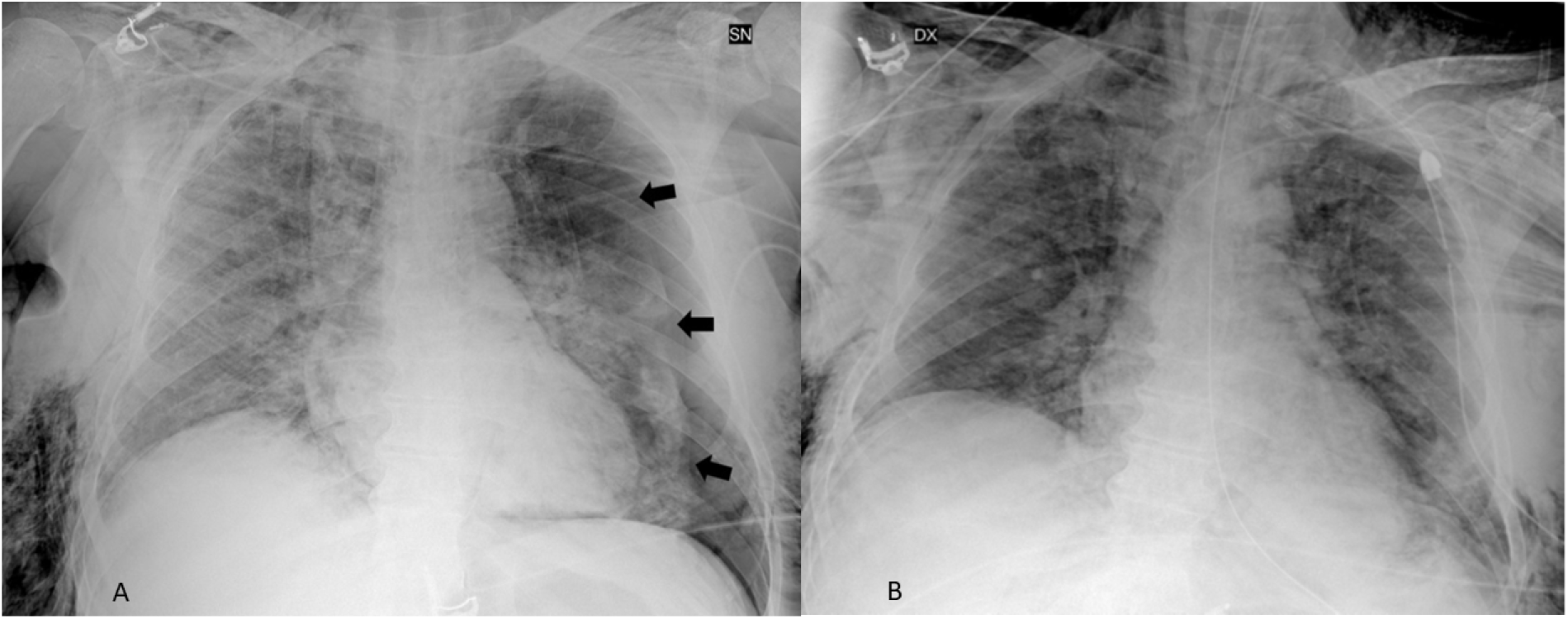
Example of PNX as visualized at Chest X-Ray (Case 3; left PNX, arrows) before (A) and after (B) drainage and ETI.

High-PEEP (>10 cmH2O) was administered to 42 (27.3%) patients, and Low-PEEP (≤10 cmH2O) to 112 (72.7%). As shown in Table 1, no statistical differences in term of characteristics were found comparing High-PEEP vs Low-PEEP group. The PNX/PNM occurred only in the High-PEEP group (5/42 [12%] vs 0/112 [0%], p<0.001).

## DISCUSSION

In our retrospective observational study PNX/PMN occurred in 3.2% of patients treated with Helmet CPAP; this incidence is higher than that reported in literature (0.95%) of all SARS-CoV2 hospitalized patients^12^, but lower than 6% of mechanically ventilated patients^13^.

Apparently, none of the patients who had this outcome had previous risk factors for PNX/PNM (no smokers, no lung diseases, no positioning central venous catheter or thoracentesis). A similar observation was reported by Martinelli et al^12^.

The incidence of PNX/PNM in non-invasive ventilated patients in our study is higher than the value reported by Martinelli et al^12^ (5/1016 [0,49%] vs 4/6574 [0.06%]), although in their study the exact number of patients treated with Helmet CPAP was not disclosed. Another notable difference between the two studies is the incidence of PNX in non-ventilated patients: no spontaneous PNX (0/1016 [0%]) were recorded by us, while a small number of cases in UK (12/6574 [0,18%]) were recorded by Martinelli et al^12^. A possible reason is the widespread use in UK of high-flow nasal cannula (HFNC) instead of the Helmet CPAP, while no HFNC devices were available in our Hospital during the observational period. This mentioned device uses lower PEEP (up to 5 cmH2O)^15^ and, consequently, it can reduce the incidence of Barotrauma. It may explain the gap between our study and Martinelli’s et al.^12^.

Survival rate in our case series of PNX/PNM was 60% in 20.3 (± 14.4) days, comparable to Martinelli et al^12^ data (PNX 63.1% PNM 53.0%) and Mirò et al^16^ (67.5%). Among these cases, the incidence of negative outcome (mortality and ETI) is relevant (40%). Moreover, even in the survived patients, PNX/PNM were associated with worsening of clinical conditions with a higher need of FiO2 (50-60% before vs 100% after). In agreement with our results, Mirò et al^16^ recently demonstrated that SARS-CoV2 patients with spontaneous PNX had a bad prognosis, even adjusting for age and sex, compared to SARS-CoV2 without PNX (OR 4.14 95% IC 1.79-9.74). Conversely, Martinelli et al^12^ reported that PNX is not an independent marker of poor prognosis; although an important limitation of their study may be the short follow up (only 28 days after the event), especially considering that 49% of patients were still hospitalized at the end of the observational period.

In our study, PNX and PNM occurred only in the High-PEEP group (11.9%). Considering the homogeneity of these groups in terms of clinical and biochemical features, we speculate that elevated PEEP may represent a risk factor for PNX/PNM in SARS-CoV2 patients. A retrospective study on SARS-CoV2 patients invasively ventilated by Kangas-Dick A. et al^17^, hypothesized a role of elevated PEEP values in generating PNX/PNM (PEEP values of 12 – 13 cmH2O). On the other hand, our cohort duration of CPAP treatment was quite variable and does not seem to play a determinant role as risk factor for events of PNX/PNM.

A possible effect of barotrauma has to be considered as superimposed to the direct effect of lung damage caused by SARS-CoV2 pneumonia. Moreover, PNX and PNM occurred rather frequently in SARS and MERS patients and some studies addressing evolution of lung lesions on CT imaging showed that in many cases spontaneous PNM occurred when ground glass opacity and consolidations began to resolve^11,18^. The pathogenesis of this phenomenon has been interpreted as the effect of the peribronchiolar inflammatory nodule formation, leading to interstitial pulmonary emphysema, tracking back along the broncho-vascular sheath and reaching the mediastinum^18^. On the other hand, histologic findings (alveolar damage with pulmonary oedema and hyaline membrane formation) appear to support the hypothesis that severe pulmonary injury predisposes the patient to spontaneous pneumothorax^19^. The diffuse alveolar damage may give rise to dilated cystic air spaces and honeycombing, predisposing the lung to air leakage from the rupture of the cystic lesions, with the consequent development of a pneumothorax. Possible shared mechanisms may underly PNX/PNM in patients with SARS-CoV2 pneumonia considering the presence of similar lung damages and imaging features as described for SARS^20,21^.

In our study PNX/PNM happened after a long time from onset of symptoms (average of 18 days) resembling what previously described in SARS patients^10^ and suggesting that a sustained period of lung inflammation might be required. Conversely, we did not find significant differences in the biochemical elements of disease severity (peak LDH and neutrophil count) that seemed to be predictable of spontaneous pneumothorax in SARS^10^ and SARS-CoV2 patients^16^. As possible explanation we reason that patients treated with Helmet CPAP were all affected by serious lung disease, so much so that we did not find any significant differences in biochemical features, while another study reported an increase in biochemical markers of severity between patients who experimented PNX^16^, but without adjusting for illness severity. Finally, in the past SARS epidemic a report conjectured that steroid therapy may play a role in the pathogenesis of spontaneous pneumothorax because it could delay wound healing and perpetuate air leakage^10^. In our study only 2 out of 5 patients have been treated with steroid therapy, so we can not speculate about this hypothesis.

Our study has several limitations. First of all, the number of events is low and for this reason our results, and in particular the association of PNX/PNM with higher PEEP, have to be confirmed in multicentric studies with a wider population. Moreover, our study is retrospective: PEEP values were decided by the clinician for each patient and in particular higher PEEP values have been set in the first weeks and subsequently lowered on the basis of clinical experience and data emerging from literature. In the end, we did not collected data about pneumoperitoneum or subcutaneous emphysema which can be classified as possible barotrauma, even if rare.

## CONCLUSION

In conclusion, our study indicates that incidence of PNX/PNM is lower in SARS-CoV2 pneumonia patients than SARS and MERS^9^. Nevertheless, the occurrence of these events worsens the prognosis. Considering the association of high PEEP (>10 cmH2O) with PNX/PNM, the use of low PEEP values has to be taken into consideration.

## Data Availability

Data are available upon reasonable request

